# Hepatitis B virus prevalence and transmission in the households of pregnant women in Kinshasa, Democratic Republic of Congo

**DOI:** 10.1101/2023.11.27.23298863

**Authors:** Camille E. Morgan, Patrick Ngimbi, Alix JN Boisson-Walsh, Sarah Ntambua, Jolie Matondo, Martine Tabala, Melchior Mwandaglirwa Kashamuka, Michael Emch, Jessie K. Edwards, Kimberly A. Powers, Linda James, Nana Mbonze, Samuel Mampunza, Marcel Yotebieng, Peyton Thompson, Jonathan B. Parr

## Abstract

**Background:** Despite routine infant vaccination and blood donor screening, the Democratic Republic of Congo (DRC) has high hepatitis B virus (HBV) prevalence compared to the United States and Europe. Through the cross-sectional Horizontal and Vertical Transmission of Hepatitis B (HOVER-HBV) study, we characterized household prevalence in DRC’s capital, Kinshasa, to inform additional prevention efforts.

**Methods:** We introduced HBV surface antigen (HBsAg) screening alongside existing HIV screening as part of routine antenatal care (ANC) in high-volume maternity clinics in Kinshasa. We recruited households of pregnant women who were HBsAg-positive and HBsAg-negative, defining households as “exposed” and “unexposed,” respectively. Household members underwent HBsAg testing and an epidemiological survey. We evaluated HBsAg prevalence and potential transmission correlates.

**Results:** We enrolled 1,006 participants from 200 households (100 exposed, 100 unexposed) across Kinshasa. HBsAg prevalence was more than twice as high in exposed households (5.0%; 95% CI: 2.8%-7.1%) as in unexposed households (1.9%; 0.6%-3.2%). Exposed direct offspring had 3.3 (0.9, 11.8) times the prevalence of unexposed direct offspring. Factors associated with HBsAg-positivity included older age, marriage, and having multiple recent partners or any new sexual partners among index mothers; and older age, lower household wealth, sharing nail clippers, and using street salons among exposed offspring.

**Conclusions:** Vertical and horizontal HBV transmission within households is ongoing in Kinshasa. Factors associated with infection reveal opportunities for HBV prevention efforts, including perinatal prevention, protection during sexual contact, and sanitation of shared personal items.

## Introduction

Despite an effective vaccine, hepatitis B virus (HBV) infection remains highly prevalent (∼6%) in Asia and Africa, resulting in significant global morbidity and mortality.^1,2^ Without a therapeutic cure, infection prevention remains the primary strategy to reduce HBV morbidity and mortality. However, HBV vaccination alone will not achieve elimination by the 2030 target.^3–5^ Modeling studies indicate that test– and-treat interventions can yield marked HBV prevalence reductions in Africa, but studies of prevention options are hindered by the limited epidemiological data from the region.^4^ While perinatal transmission is the dominant driver of ongoing endemicity in Asia,^6–8^ available studies suggest a greater contribution of household and community (“horizontal”) transmission in Africa, particularly during early childhood^9,10^ but also in adulthood.^4^ Improved understanding of dominant HBV transmission modes and risk factors is needed to design effective interventions in Africa, especially in HBV-endemic countries like the Democratic Republic of Congo (DRC).

National HBV prevalence in the DRC is estimated to be 3.3% by HBV surface antigen (HBsAg) testing,^11^ translating to approximately 2.5 million chronic infections in a country where advanced hepatology care is essentially inaccessible.^12^ Estimated prevalence is higher among blood donors,^13^ women with HIV presenting to urban antenatal care (ANC) settings,^14^ pregnant women in rural areas,^15,16^ healthcare workers,^17^ and survivors of sexual violence.^18^ Blood donor screening and the three-dose infant pentavalent vaccine series are the only HBV prevention measures implemented nationally in DRC, but complete infant HBV vaccination coverage is <70%.^19^ For prevention of mother-to-child transmission, the World Health Organization recommends ANC HBsAg screening, maternal antiviral prophylaxis, and infant birth-dose vaccination to prevent perinatal transmission.^20^ These activities are feasible in the DRC but not yet implemented.^21,22^.

To investigate HBV correlates and inform expanded interventions in the DRC, we conducted the Horizontal and Vertical Transmission of Hepatitis B (HOVER-HBV) study. We built upon the established prevention of mother-to-child HIV transmission (PMTCT) program infrastructure to introduce ANC HBsAg screening, characterize HBV prevalence in ANC patients’ households across urban Kinshasa, and identify attributes and practices associated with HBsAg-positivity.

## Methods

### Study design and participant recruitment

To recruit households, we introduced DETERMINE 2^23^ (Abbott, Abbott Park, IL) point-of-care (POC) HBsAg testing alongside existing ANC HIV testing in high-volume maternity centers in Kinshasa (**Figure 1**). During two recruitment periods, pregnant women screened for HBsAg were offered enrollment of their households in the study. In this matriarchal design, recruited women served as “index mothers” for enrolled households, with a pre-specified target of 100 households of HBsAg-positive mothers and 100 households of HBsAg-negative mothers.

**Figure 1.**
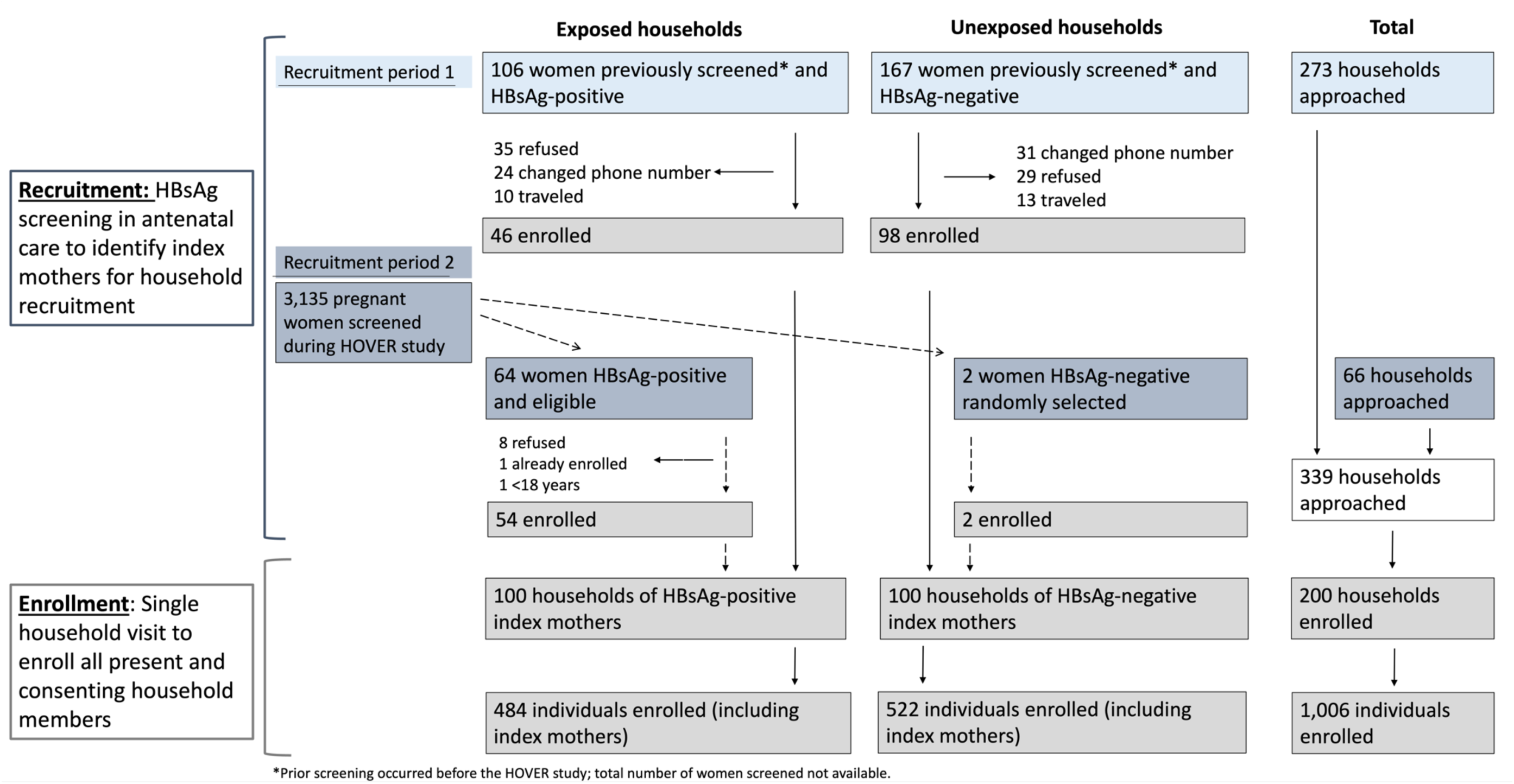
Recruitment and enrollment of households of HBsAg-positive and HBsAg-negative index mothers.

### Study procedures

Data collection at the single household visit included POC HBsAg testing (including repeat testing of the index mothers), collection of dried blood spot (DBS) specimens, and administration of household and individual questionnaires (**Appendix 1**) covering demographics and potential sources of horizontal HBV transmission within household and community settings. HIV infection and antiretroviral therapy (ART) use, particularly tenofovir-based regimens given its anti-HBV activity, were determined by self-report. As HBV can remain infectious on surfaces for at least 7 days,^24^ we collected information about household and community practices that could result in HBV transmission, based on past findings from other countries in the region.^16,25–27^ We offered the three-dose Euvax-B HBV (LG Life Sciences, Republic of Korea) vaccination to all HBsAg-negative individuals living with someone who was HBsAg-positive, and recorded all resulting vaccinations occurring at the two centers where we offered them up to six months after the last enrollment.

### Analytical approach

We defined HBV-“exposed” and “unexposed” households as households of index mothers who were HBsAg-positive and HBsAg-negative during ANC recruitment, respectively; in sensitivity analyses, we applied alternate definitions of household exposure based on index mothers’ HBsAg status across the recruitment and enrollment timepoints (**Supplementary Material**). We conducted descriptive analyses of household composition, HBsAg positivity patterns within households, and household/participant demographics, including composite indices of modern housing and wealth to approximate standard of living across households (**Supplementary Material**). We considered participant age both continuously and categorically, defining categories based on age relative to the introduction of the three-dose HBV vaccination in the national infant immunization program (**Supplementary Material**).^28,29^

In our primary analysis, we compared HBsAg prevalence between exposed and unexposed households and between household member types (offspring versus other). We estimated prevalence ratios for each comparison, first unadjusted and then adjusted for household clustering using a random intercept for household. To examine potential correlates of HBV infection, we also estimated measures of association between HBsAg positivity and individual, household, and community-level attributes and practices. Variable coding is detailed in Supplementary Material. Briefly, individual attributes included age and marital status; household variables included household wealth, sharing personal objects within the household, and premasticating food for another household member; community variables included receiving blood transfusions, manicures/pedicures, tattoos, traditional scarification, and a variety of sexual behaviors. Given the case-control design of index mother recruitment, we used logistic regression to estimate odds ratios for these factors’ associations with maternal HBsAg positivity. For the analysis among household members, we conducted stratified analyses by household member type (offspring vs. other household member), estimating a prevalence ratio for each attribute/practice from multilevel log-binomial regression with a random intercept to account for household clustering. Where log-binomial models failed to converge, we used the odds ratio from the logistic regression model to approximate the prevalence ratio, which is minimally biased for rare outcomes (<10% prevalence) such as HBsAg positivity.^30^ We calculated 95% confidence intervals (CI) to assess precision of each estimate. In subgroups with fewer than 10 HBsAg infections, we calculated Fisher’s exact p-values.

All data were imported into R (v4.1.1, R Core Team, Vienna, Austria) using the *REDCapR* package (v1.1.0). We analyzed data using the *tidyverse* (v2.0.0), *tableone* (v0.13.2), and *lme4* (v1.1.32) packages. R code is publicly available at https://github.com/IDEELResearch/hbv_hover. This study was approved by the Institutional Review Board at the University of North Carolina (19-1875) and the Ethics Committee at Université Protestante au Congo (CE/UPC/0062).

## Results

### Study population

From February 2021 to September 2022, we offered enrollment to 339 households, and enrolled 200 households and 1,006 individuals (**Figure 1**). Overall, 190 index mothers were recruited from two maternity centers, and 10 women from nine other maternity centers. Participating households were located in neighborhoods across metropolitan Kinshasa (**Figure 2A**). Few households (20.5%) lived in modern housing structures, and most participants were transient (median of two years [IQR: 1, 5] in the home) (**Table 1**). We enrolled a median of 5 (IQR: 3, 6) members per exposed and unexposed household (**Figure 2B**). Most recruited index mothers were multiparous: in 176 (88%) households, we enrolled at least one direct offspring, with a median of 3 children (IQR: 1,4) enrolled in both exposed and unexposed households (**Table 1**). In 86 households (43%), we enrolled the index mother’s sexual partner, with partners enrolling in a considerably higher proportion of exposed (52%) than unexposed (34%) households. Index mothers had a median age of 32 years (IQR: 27, 37), with a higher median age among mothers in exposed (33 years) versus unexposed households (30 years). We enrolled 467 direct offspring of index mothers (228 exposed, 239 unexposed), with similar age distributions in exposed and unexposed households. Most offspring (82% exposed, 84% unexposed) were 13 years of age or younger in 2022, and thus born after three-dose infant HBV vaccination was introduced in DRC. We enrolled 331 other household members, with the most common relationships to index mothers being nieces/nephews (n=93), siblings (n=90), and partners (n=86).

**Figure 2.**
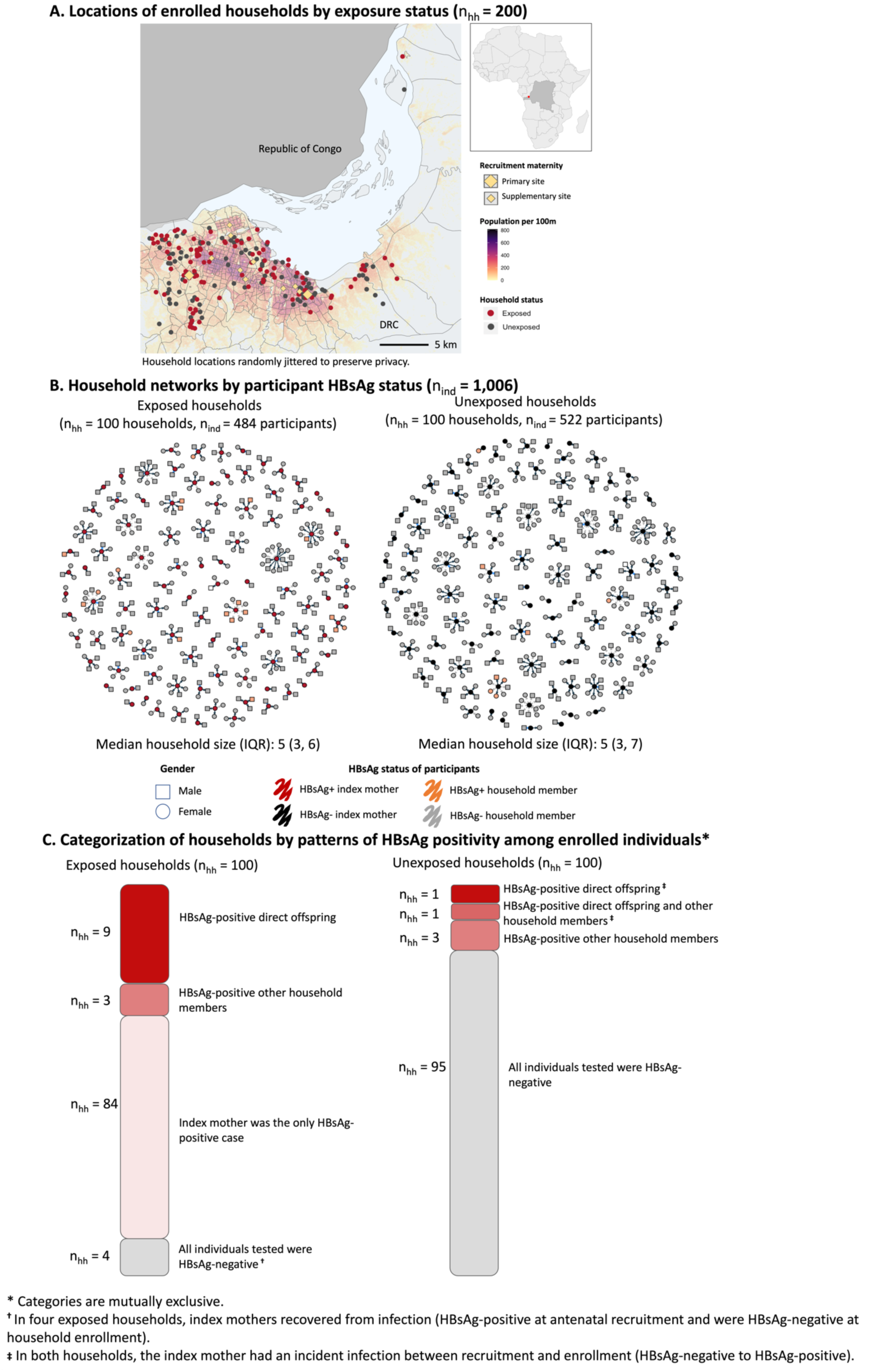
Enrolled households by geography and household structure.

**Table 1.** Characteristics of study households (A) and participants (B) enrolled in the HOVER-HBV study.

| Characteristic | Exposed households<br>(HBsAg+ index mother)* | Unexposed households<br>(HBsAg- index mother)* | Overall |
| --- | --- | --- | --- |
| n | 100 | 100 | 200 |
| Household members enrolled, median (IQR) | 5 (3, 6) | 5 (3, 7) | 5 (3, 6) |
| Enrolled direct offspring of index mothers per household, median (IQR) | 3 (1, 4) | 3 (1, 4) | 3 (1, 4) |
| Household structures |  |  |  |
| Vertical: Index mother/direct offspring | 87 (87%) | 89 (89%) | 176 (88%) |
| Sexual: index mother and male partner enrolled | 52 (52%) | 34 (34%) | 86 (43%) |
| Modern housing (%)<sup>†</sup> | 21 (21%) | 20 (20%) | 41 (20.5%) |
| Wealth quartile (%)<sup>‡</sup> |  |  |  |
| Lowest | 28 (28%) | 22 (22%) | 50 (25%) |
| Lower middle | 20 (20%) | 30 (30%) | 50 (25%) |
| Upper middle | 24 (24%) | 26 (26%) | 50 (25%) |
| Highest wealth | 28 (28%) | 22 (22%) | 50 (25%) |

**B. Individuals**
| Characteristic | Index mothers |  | Direct offspring |  | Other household members |  | Overall |
| --- | --- | --- | --- | --- | --- | --- | --- |
|  | Exposed | Unexposed | Exposed | Unexposed | Exposed | Unexposed |  |
| n | 100 | 100 | 228 | 239 | 156 | 183 | 1,006 |
| HBsAg testing, n | 100 | 100 | 227 | 237 | 156 | 183 | 1,003 |
| HBsAg+ at enrollment, n (%) | 96 (96.0) | 2 (2.0) | 12 (5.3) | 3 (1.3) | 7 (4.5) | 5 (2.9) | 125 |
| Age, median years (IQR) | 33 (29, 37) | 30 (25, 37) | 6 (2.6, 11) | 6 (2, 11) | 26 (13, 40) | 22 (13.2, 38) | 15 (5, 30) |
| Age group, years (%)<sup>§</sup> |  |  |  |  |  |  |  |
| >13 | 100 (100.0) | 100 (100.0) | 41 (18.0) | 38 (15.9) | 121 (77.6) | 145 (79.2) | 545 (54.2) |
| ≤13 | 0 (0.0) | 0 (0.0) | 187 (82.0) | 201 (84.1) | 35 (22.4) | 38 (20.8) | 461 (45.8) |
| Female sex, n (%) | 100 (100.0) | 100 (100.0) | 119 (52.5) | 142 (59.5) | 76 (48.1) | 95 (55.2) | 637 (63.3) |
| Relationship to index mother, n (%) |  |  |  |  |  |  |  |
| Index mother | 100 (100.0) | 100 (100.0) | 0 (0.0) | 0 (0.0) | 0 (0.0) | 0 (0.0) | 200 (19.9) |
| Son/daughter | 0 (0.0) | 0 (0.0) | 228 (100.0) | 239 (100.0) | 0 (0.0) | 0 (0.0) | 475 (46.4) |
| Current male partner | 0 (0.0) | 0 (0.0) | 0 (0.0) | 0 (0.0) | 52 (33.1) | 34 (18.6) | 86 (8.5) |
| Brother/sister | 0 (0.0) | 0 (0.0) | 0 (0.0) | 0 (0.0) | 39 (24.8) | 60 (32.8) | 90 (9.9) |
| Nephew/niece | 0 (0.0) | 0 (0.0) | 0 (0.0) | 0 (0.0) | 41 (26.1) | 53 (29.0) | 93 (9.3) |
| Other | 0 (0.0) | 0 (0.0) | 0 (0.0) | 0 (0.0) | 25 (15.9) | 36 (19.7) | 61 (6.0) |
| Marital status, n (%) |  |  |  |  |  |  |  |
| Married or living with someone | 85 (85.0) | 75 (75.0) | 0 (0.0) | 2 (7.4) | 56 (50.0) | 49 (37.4) | 267 (52.8) |
| Never married | 7 (7.0) | 16 (16.0) | 36 (100.0) | 25 (92.6) | 45 (40.2) | 66 (50.4) | 195 (38.5) |
| Divorced/widowed | 8 (8.0) | 9 (9.0) | 0 (0.0) | 0 (0.0) | 11 (9.8) | 16 (12.2) | 44 (8.7) |
| Household is primary residence, n (%) | 98 (98.0) | 98 (98.0) | 225 (98.7) | 234 (98.3) | 135 (86.5) | 163 (89.1) | 954 (94.8) |
| Slept in household last night, n (%) | 100 (100.0) | 99 (99.0) | 225 (99.1) | 238 (99.6) | 144 (92.9) | 178 (97.2) | 984 (98.0) |
| ≥1 past positive HBV test, n (%) | 90 (90.0) | 0 (0.0) | 0 (0.0) | 1 (0.4) | 3 (1.9) | 1 (0.5) | 95 (9.4) |
| Past positive HIV test, n (%) | 16 (16.0) | 1 (1.0) | 3 (1.3) | 0 (0.0) | 1 (0.6) | 1 (0.5) | 22 (2.2) |
| Current HAART regimen, n (%) |  |  |  |  |  |  |  |
| Tenofovir, lamivudine, dolutegravir | 15 (93.8) | 1 (100.0) | 1 (33.3) | 0 (0.0) | 1 (100.0) | 1 (100.0) | 19 (86.4) |
| Dolutegravir, abacavir | 0 (0.0) | 0 (0.0) | 2 (66.7) | 0 (0.0) | 0 (0.0) | 0 (0.0) | 2 (9.1) |
| Not taking | 1 (6.2) | 0 (0.0) | 0 (0.0) | 0 (0.0) | 0 (0.0) | 0 (0.0) | 1 (4.5) |
| Pregnant at household enrollment, n (%) | 56 (56.6) | 13 (13.0) | 0 (0.0) | 0 (0.0) | 0 (0.0) | 3 (4.4) | 72 (20.2) |

\*Exposure status based on index mother's antenatal recruitment point-of-care HBsAg screening.
†Modern housing is a composite variable of materials used in the roofing, walls, flooring, and windows, using previously published categorizations further described in the Supplementary Material.
\*Wealth index is calculated by a principal components analysis accounting for household attributes further described in the Supplementary Material.
§Age in 2022 dichotomized using 13 years to approximate whether participant was born before or after infant pentavalent vaccination introduction in 2009, further described in Supplementary Material.
¶Survey question asked to those age 15 years and above.

### HBsAg prevalence

HBsAg prevalence among index mothers’ household members was 5.0% (95% CI: 2.8%, 7.1%) and 1.9% (0.6%, 3.2%) in exposed and unexposed households, respectively, corresponding to an unadjusted prevalence ratio (PR) of 2.61 (1.20, 6.25), and PR adjusted for household clustering of 2.52 (0.88, 7.23) (**Table 2, Supplementary** Figure 3). Overall, we observed 27 household members who were HBsAg-positive (19 exposed, 8 unexposed) from 17 distinct households (**Figures 2B-2C**). We observed 15 HBsAg infections among direct offspring; HBsAg prevalence among direct offspring was 5.3% (2.4%, 8.2%) in exposed households and 1.3% (0.0%, 2.7%) in unexposed households, corresponding to an unadjusted PR of 4.18 (1.35, 18.16) and adjusted PR of 3.33 (0.94, 11.84). Among these 15 direct offspring who were HBsAg-positive, 12 had mothers who were HBsAg-positive at both timepoints; the remaining three came from two households in which index mothers had incident infections. We observed 12 HBsAg infections among other household members; HBsAg prevalence was 4.5% (1.2%, 7.7%) and 2.7% (0.4%, 5.1%) among other household members in exposed and unexposed households, respectively, corresponding to an unadjusted PR of 1.64 (95%CI: 0.53, 5.45) and adjusted PR of 1.01 (95% CI: 0.24, 4.25). We observed one HBsAg infection among exposed male partners and one among unexposed male partners, for a PR of 0.65 (95% CI: 0.04, 10.10) comparing those exposed to those unexposed.

**Table 2.**
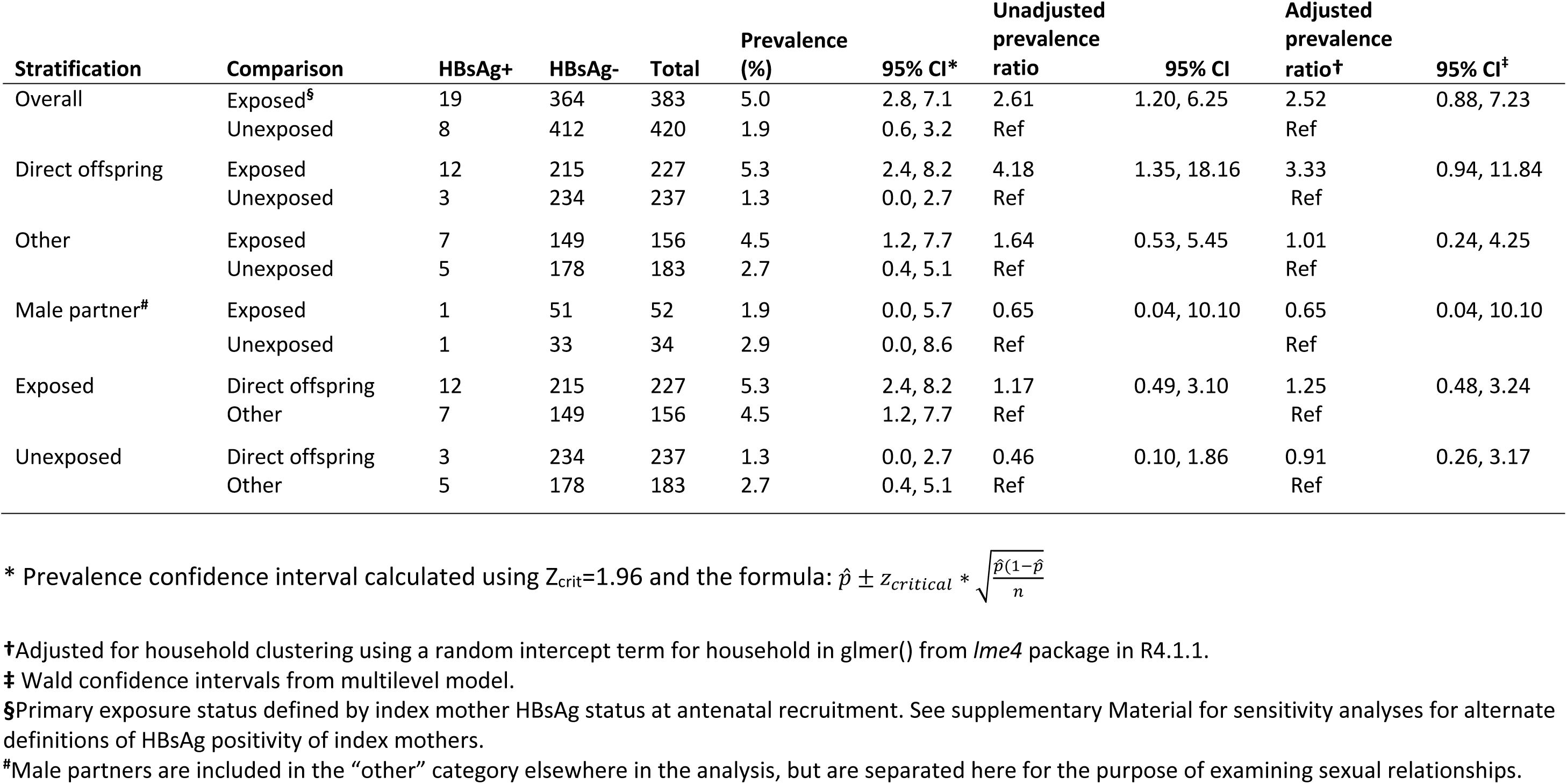
HBsAg prevalence among exposed and unexposed household members.

| Stratification | Comparison | HBsAg+ | HBsAg- | Total | Prevalence (%) | 95% CI* | Unadjusted prevalence ratio | 95% CI | Adjusted prevalence ratio† | 95% CI‡ |
| --- | --- | --- | --- | --- | --- | --- | --- | --- | --- | --- |
| Overall | Exposed§ | 19 | 364 | 383 | 5.0 | 2.8, 7.1 | 2.61 | 1.20, 6.25 | 2.52 | 0.88, 7.23 |
|  | Unexposed | 8 | 412 | 420 | 1.9 | 0.6, 3.2 | Ref |  | Ref |  |
| Direct offspring | Exposed | 12 | 215 | 227 | 5.3 | 2.4, 8.2 | 4.18 | 1.35, 18.16 | 3.33 | 0.94, 11.84 |
|  | Unexposed | 3 | 234 | 237 | 1.3 | 0.0, 2.7 | Ref |  | Ref |  |
| Other | Exposed | 7 | 149 | 156 | 4.5 | 1.2, 7.7 | 1.64 | 0.53, 5.45 | 1.01 | 0.24, 4.25 |
|  | Unexposed | 5 | 178 | 183 | 2.7 | 0.4, 5.1 | Ref |  | Ref |  |
| Male partner# | Exposed | 1 | 51 | 52 | 1.9 | 0.0, 5.7 | 0.65 | 0.04, 10.10 | 0.65 | 0.04, 10.10 |
|  | Unexposed | 1 | 33 | 34 | 2.9 | 0.0, 8.6 | Ref |  | Ref |  |
| Exposed | Direct offspring | 12 | 215 | 227 | 5.3 | 2.4, 8.2 | 1.17 | 0.49, 3.10 | 1.25 | 0.48, 3.24 |
|  | Other | 7 | 149 | 156 | 4.5 | 1.2, 7.7 | Ref |  | Ref |  |
| Unexposed | Direct offspring | 3 | 234 | 237 | 1.3 | 0.0, 2.7 | 0.46 | 0.10, 1.86 | 0.91 | 0.26, 3.17 |
|  | Other | 5 | 178 | 183 | 2.7 | 0.4, 5.1 | Ref |  | Ref |  |

\* Prevalence confidence interval calculated using $Z_{crit}=1.96$ and the formula: $\hat{p} \pm Z_{critical} * \sqrt{\frac{\hat{p}(1-\hat{p})}{n}}$
†Adjusted for household clustering using a random intercept term for household in glmer() from lme4 package in R4.1.1.
‡ Wald confidence intervals from multilevel model.
§Primary exposure status defined by index mother HBsAg status at antenatal recruitment. See supplementary Material for sensitivity analyses for alternate definitions of HBsAg positivity of index mothers.
#Male partners are included in the “other” category elsewhere in the analysis, but are separated here for the purpose of examining sexual relationships.

In sensitivity analyses using alternate definitions of HBV exposure, household HBsAg prevalence estimates were largely consistent with those obtained under the primary exposure definition (Supplementary Figure 3), except for the analysis of direct offspring (all direct offspring infected lived with a mother who was HBsAg-positive at least once). Of the 100 index mothers who were HBsAg-positive at recruitment, 96 were HBsAg-positive and four were HBsAg-negative at enrollment (**Figure 2C**). Two women who were HBsAg-negative at recruitment were HBsAg-positive at enrollment, representing incident cases of horizontal transmission, and 98 index mothers were HBsAg-negative at both points.

### Factors associated with HBsAg positivity

Among index mothers, several individual attributes and potential sources of community HBV exposure were associated with HBsAg positivity. Increasing age was associated with higher odds of HBsAg positivity, with an odds ratio of 1.06 (1.01, 1.11) per one-year increase in age (**Figure 3, Supplementary** Figure 4). Never having been married was associated with 0.39 (95% CI: 0.14, 0.96) times the odds of HBsAg positivity compared with being married. Declining to answer age of sexual debut was associated with 4.53 (95% CI: 1.81, 12.79) times the odds of HBsAg positivity compared with reporting sexual debut at 18 years or older; sexual debut before 18 years was associated with higher odds of HBsAg positivity compared with ≥18 years, but had a lower magnitude of association than declining to answer (OR: 1.21, 95% CI 0.66, 2.23). All variables assessing recent multiple and new sexual partners were associated with higher odds of HBsAg positivity compared with the referent; for example, having at least one new sexual partner in the last three months or declining to answer was associated with 3.63 (95% CI: 1.60, 9.06) times the odds of HBsAg positivity compared with having no new sexual partners in the last three months. All associations held across sensitivity analyses in which definitions of household HBV exposure status were varied (**Supplementary** Figure 4). We did not observe evidence that engaging in transactional sex was associated with higher odds of HBsAg positivity (OR: 1.00, 95% CI: 0.53, 1.89).

**Figure 3.**
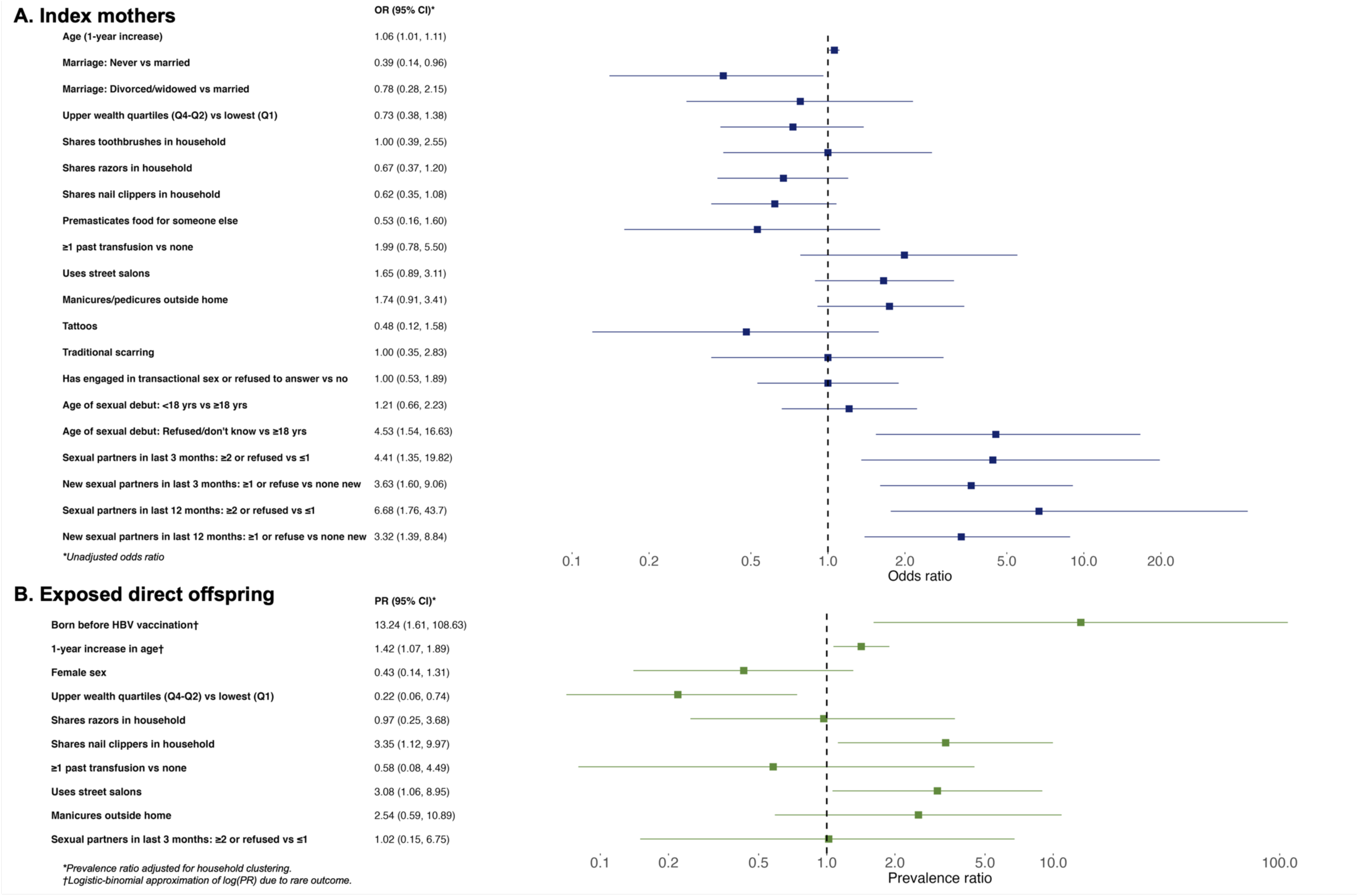
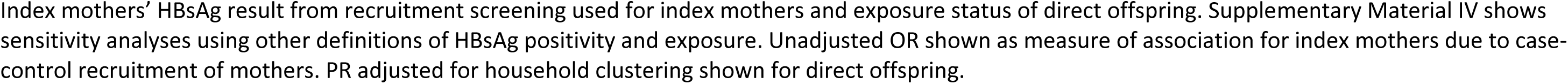
Attributes and practices associated with HBsAg-positivity among index mothers (A) and exposed direct offspring (B).

Among exposed direct offspring, a one-year increase in age was associated with higher HBsAg prevalence [adjusted PR = 1.42; 95% CI = (1.07,1.89)]. Offspring older than 13 years in 2022 (born before pentavalent vaccine introduction in DRC) had 13.24 (1.61, 108.63) times the HBsAg prevalence of those ≤13 years (**Figure 3, Supplementary** Figure 5). Exposed offspring from wealthier households had 0.22 [95% CI: (0.06, 0.74)] times the HBsAg prevalence of those from households in the lowest wealth quartile. Sharing nail clippers in the household [adjusted PR: 3.35; 95% CI: (1.12, 9.97)] and using street salons [adjusted PR: 3.08; 95% CI: (1.06, 10.89)] in the community were both associated with higher HBsAg prevalence among exposed direct offspring. No attributes or practices were associated with HBsAg-positivity among unexposed direct offspring, but this analysis was limited by few (n=3) unexposed direct offspring who were HBsAg-positive (**Supplementary Table 3**).

Exposed and unexposed other household members had too few infections (n=7 and n=5, respectively) to estimate PRs within these subgroups, but among exposed other household members, history of traditional scarring was associated with HBsAg-positivity (p-value = 0.002; **Supplementary Table 4**).

Among unexposed other household members, no attributes or practices were significantly associated with HBsAg positivity (**Supplementary Table 5**).

### HBV vaccination

A total of 330 HBsAg-negative household members were living with someone who was HBsAg-positive. At six months following enrollment completion, 162 (49%) had initiated the vaccine series and 149 (45%) had completed the series (92% completion rate); 51 (15%) had refused vaccination outright; the remainder had accepted but did not present for vaccination. When we followed up with vaccine-eligible participants, the most-cited reasons for non-vaccination were vaccine hesitancy and distance to the maternities where we offered vaccination.

## Discussion

In this large household HBV investigation in the DRC, we identified evidence of ongoing HBV transmission and opportunities for a range of HBV prevention efforts. HBsAg prevalence was higher in the households of HBsAg-positive mothers, indicating that the existing HIV PMTCT infrastructure in countries like the DRC could be used to identify households for targeted prevention. Factors associated with HBsAg-positivity among index mothers included increasing age, current marriage, and recent sexual behavior (having at least two recent partners, having at least one new recent partner, or declining to answer recent partner questions). Among exposed direct offspring, HBsAg-positivity was associated with increasing age, fewer household resources, and use of shared nail clippers or street salons. We also observed evidence that traditional scarring could be associated with HBsAg positivity among other household members. These potential sources of HBV infection corroborate findings from past studies in other African settings,^25,31^ and suggest priority behaviors or subgroups for intervention in urban Kinshasa that may be relevant to other megacities in Africa.

Roll-out of infant HBV vaccination within the pentavalent series starting at six weeks of age is one possible explanation for the HBV prevalence among exposed children, which was more than 13 times as high for those who were >13 years compared to those who were ≤13 years in 2022. Our findings are consistent with results of a study conducted in Burkina Faso, which also reported lower HBsAg positivity among children born to 215 HBsAg-positive mothers after HBV vaccine rollout but before birth dose.^32^ We observed relatively few HBsAg-positive offspring, suggestive of infrequent perinatal transmission given that proven PMTCT measures have not been implemented in DRC. Our observation of incident and cleared infections among mothers further suggests recent HBV exposures and that horizontal transmission is occurring. Accumulation of HBV exposures in households and communities over time is a plausible explanation for the observed increasing HBsAg-positivity risk with age.

The strong observed associations with sexual exposures provides further evidence of horizontal HBV transmission. The strongest association with HBsAg positivity was observed for index mothers who reported multiple recent or any new sexual partners or who declined to discuss. This finding is in line with a past study of DRC healthcare workers that found an association between multiple sexual partners and HBsAg positivity.^17^ We did not observe evidence that engagement in transactional sex was associated with HBsAg positivity. Interestingly, we observed lower HBsAg prevalence among male partners of HBsAg-positive mothers compared with HBsAg-negative mothers. While precision was limited, one explanation for this finding is that sexual partners have been exposed and recovered from infection, which could be clarified by serological analysis that was not feasible with DBS samples. Together, our findings indicate that development of HBV prevention efforts for the broader population of women of childbearing age are needed in the DRC.

Our experience in this study indicates that integration of antenatal HBsAg testing alongside existing HIV testing is acceptable to maternity center staff and patients, consistent with prior findings.^21^ While rapid HBsAg tests are available for just over 1 USD per test, scaling this effort is often hindered by a siloed approach to healthcare in which HIV funders do not cover HBsAg or syphilis screening, both of which are recommended for triple elimination.^33^ Women who are HBsAg-positive could be offered HBV antiviral prophylaxis and HBV birth-dose vaccine for their newborns, and given the opportunity to have household members screened and vaccinated. ANC visits also provide opportunities for health education. While these measures are not currently offered in the DRC, increasing evidence indicates that they would be effective and feasible in the DRC.^21^

Our study has several limitations. First, our cross-sectional study design does not allow for analysis of the timing of infection, preventing definitive determination of vertical vs. horizontal transmission. Serological analysis and HBV sequencing could improve our characterization of these transmission patterns, but serological assays for DBS samples collected in this study and often used in resource-limited settings like DRC continue to perform poorly for HBV.^34^ Second, enrollment of households several months to over a year after recruitment during antenatal screening meant that we inherently selected a population based in Kinshasa. Individuals with frequent time out of Kinshasa might have a different prevalence of HBV infection and associated behaviors not captured in this study. Third, household members absent during our enrollment visit and thus not included in the study may also be infected, potentially resulting in biased prevalence estimates. However, our estimates remain useful for clinicians assessing household infection risk as part of routine antenatal care and for development of targeted prevention programs.

## Conclusions

In the largest and most detailed household investigation of HBV in DRC conducted to-date, we found that HBV screening as part of existing HIV PMTCT programs can be used to identify infected mothers and household members, and households where vaccination may be particulary beneficial. In addition to WHO-recommended efforts to prevent mother-to-infant HBV transmission, prevention of horizontal transmission within households and communities should be prioritized. Possible interventions include education to reduce blood exposure through household item sharing and to improve protection during sexual intercourse in affected households, as well as targeted vaccination programs in adults. While additional research is needed to determine precise HBV transmission mechanisms in settings like the DRC, our findings provide a foundation for developing new HBV transmission prevention strategies.

## Data Availability

All data produced in the present study are available upon reasonable request to the authors.

## Acknowledgements

We would like to thank all participants who offered their time and participation in this study. We would also like to thank the staff at Binza and Kingasani maternities for their collaboration in conducting antenatal HBsAg screening and administering Euvax HBV vaccination. We would also like to thank Dr. Alpha Oumar Diallo for his contributions establishing the REDCap study database. Lastly, we thank the late Dr. Steve Meshnick for his mentorship and contributions to the conceptualization of this study. Data may be made available upon reasonable request. JBP, PT, MY, and SM conceptualized the study. PN, SN, JM, MT, NM, and MMK collected data. CEM, AJNB, PN, SN, JM, NM, MMK, LJ, JBP, PT, ME, JKE, and KAP analyzed and interpreted results. CEM, AJNB, JBP, PT, ME, JKE, and KAP drafted the manuscript. All authors have reviewed and approved the manuscript.

