## Supplementary Material for "Hepatitis B virus prevalence and transmission in the households of pregnant women in Kinshasa, Democratic Republic of Congo"

**Methods: Variable coding**

### I. Categorical age grouping

A categorical age variable was used to dichotomize participants over 13 years or 13 years or younger. This cutoff was used to distinguish participants who have been born since infant HBV vaccination was introduced in the national infant immunization program in DRC. Infant HBV vaccination included in the pentavalent vaccine (DTP-HepB-Hib) was introduced in 2009,[1,2] and we use participants age in 2022 as the cutoff. A tetravalent vaccine (DTP-HepB) was introduced in 2007 but had poor rollout,[3] so the analysis focuses on pentavalent vaccination.

Published dates of HBV vaccine introduction retrieved from:


1. Le gouvernement de of the Democratic Republic of Congo. Rapport annuel de situation 2008 (Français). Published 14 mai 2009. https://www.gavi.org/sites/default/files/document/annual-progress-report-congo%2C-democratic-republic-of-the-2008--francais-pdf.pdf)
2. The Government of the Democratic Republic of Congo. Annual progress report 2010 (English). Published 01 June 2011. https://www.gavi.org/sites/default/files/document/annual-progress-report-congo%2C-democratic-republic-of-the-2010pdf.pdf)
3. The Government of the Democratic Republic of Congo. Annual progress report 2007 (English). Published online April 30, 2008. https://www.gavi.org/sites/default/files/document/annual-progress-report-congo%2C-democratic-republic-of-the-2007pdf.pdf)


### II. Modern housing

Modern housing is a composite variable of materials used in the roofing, walls, flooring, and windows, using previously published categorizations from Deutsch-Feldman et al (2020). Each of these components was categorized as modern vs not modern using the divisions described below. The modern housing indicator were households in which the roofing, walls, flooring and windows were "modern".

Roofing:

- "Modern":
  - English: metal, zinc/cement, tiles/slate, or cement
  - Français: Tôle, zinc/fibre de ciment, tuiles, béton (ciment), shingles
- "Not modern":
  - English: leaves, earth, mats, bamboo, wood, cardboard/plywood
  - Français: Tôle, chaume/palme/feuilles, mottes de terre, nattes, palmes/bambou, planches en bois, carton, bois

Walls:

- "Modern":
  - English: cement, stone, bricks, covered adobe
  - Français: Zinc/fibre de ciment, tuiles, béton (ciment), shingles

- “Not modern”:
    - English: Metal, leaves, earth, mats, bamboo, wood, cardboard/plywood, uncovered adobe
    - Français: Tôle, chaume/palme/feuilles, mottes de terre, adobe non recouvert, nattes, palmes/bambou, planches en bois, carton, bois

Flooring:
- “Modern”:
    - English: vinyl, asphalt, ceramic tiles, cement, or carpet, parquet/polished wood
    - Français: Bandes de vinyle/asphalte, carrelage, ciment, moquette, parquet/bois ciré
- “Not modern”:
    - English: Earth/sand, dung, wooden boards, palms/bamboo
    - Français: Terre/sable, bouse, planches en bois, palmes/bambou

Windows:
- “Modern”:
    - English: glass, screen
    - Français: verre, rideaux de tulle
- “Not modern”:
    - English: open/no windows, plastic/paper/carton, wood/planks/metal
    - Français: ouverte/pas de fenêtres, papier en plastique/carton, bois/planche/tôle

Deutsch-Feldman M, Brazeau NF, Parr JB, et al. Spatial and epidemiological drivers of Plasmodium falciparum malaria among adults in the Democratic Republic of the Congo. BMJ Global Health 2020; 5: e002316).

### III. Wealth index

*Statistical analysis*

We used *prcomp()* in R.4.1.1 to conduct a principal component analysis (PCA) to calculate wealth quartiles as a proxy for households’ standard of living. Approximating wealth using a PCA is an imperfect measure of socioeconomic status (SES), but this method is widely accepted (1, 2). Wealth index is calculated by a principal components analysis accounting for household attributes (household electricity access, water supply, modern flooring, modern walls, modern roofing, windows, toilet, refrigerator, oven, stove, generator, television, radio, gardening hoes, sewing machines) and ownership of items by household members (computers, cars, motorbikes, bicycle, rental houses). No missing data were observed. The first component explained 15.7% of the variance and following similar studies (Supplementary Figure 1),(3) 25$^{th}$, 50$^{th}$, and 75$^{th}$ percentiles of the distribution (Supplementary Figure 2) were used to calculate wealth quartiles.

1. Howe LD, Hargreaves JR, Huttly SR. Issues in the construction of wealth indices for the measurement of socio-economic position in low-income countries. Emerg Themes Epidemiol. 2008;5:3.

2. Vyas S, Kumaranayake L. Constructing socio-economic status indices: how to use principal components analysis. Health Policy Plan. 2006;21(6):459-68.
3. Thompson P, Morgan CE, Ngimbi P, et al. Arresting vertical transmission of hepatitis B virus (AVERT-HBV) in pregnant women and their neonates in the Democratic Republic of the Congo: a feasibility study. The Lancet Global Health. Published online August 2021:S2214109X21003041. doi:10.1016/S2214-109X(21)00304-1.

**Supplementary Figure 1:** Variance explained by each principal component resulting from principal component analysis for wealth.

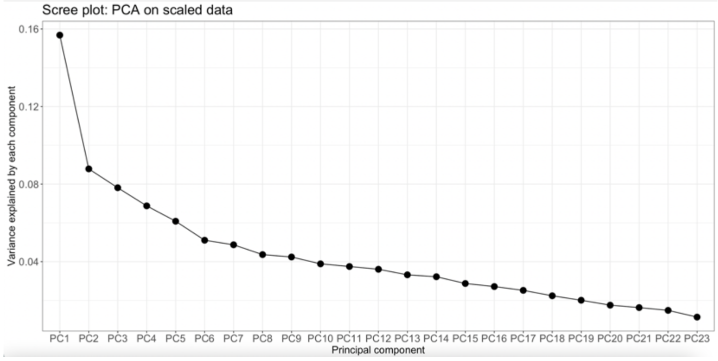


**Supplementary Figure 2:** Distribution of values for Principal Component 1, from which wealth quartiles were calculated.

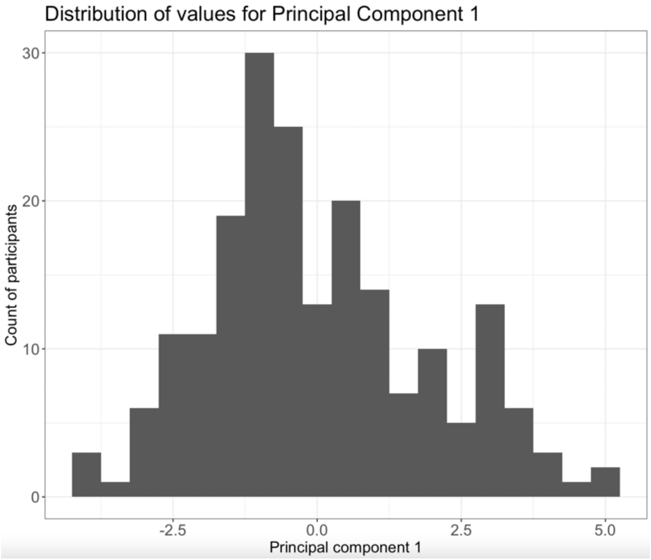

Individual, household, and community-level variables

Individual

- Age, continuous: 1-year increase in age to reflect aggregated exposures to HBV.
- Age, categorical: Based on birth prior to vs since infant pentavalent (includes HBV) vaccination was introduced in DRC. >13 years in 2022 (born prior to pentavalent introduction in 2009 and thus likely not vaccinated) vs ≤ 13 years in 2022 (referent group, born since pentavalent introduction).
- Marital status: collapsed into three groups, never married, currently married or living together, and divorced, separated, or widowed.
    - Never married vs currently married/living together (referent)
    - Divorced, separated, or widowed vs currently married/living together (referent)

Household variables

- Household wealth: categorized as upper three quartiles compared with the lowest quartile (index described above).
- Sharing of personal objects with other household members, analyzed separately:
    - Nail clippers (reports sharing with others vs doesn't share)
    - Razors (reports sharing with others vs doesn't share)
    - Toothbrushes (reports sharing with others vs doesn't share)
- Premastication of food: premasticates food or chewing gum before giving to another household member vs does not.

Community variables

- Blood transfusions: receipt of at least one blood transfusion in the past vs never received (referent)
- Manicures or pedicures outside the home: at least one/month vs never uses (referent)
- Tattoos: ever received vs never received (referent)
- Traditional scarification: yes or refused vs never received (referent)
- Age of sexual debut:
    - < 18 years vs ≥ 18 years (referent)
    - Refused to answer age of sexual debut vs ≥ 18 years (referent)
- Transactional sex: reports having given or received money in exchange for sex or refused to answer versus reports never giving or receiving money for sex (referent).
- Number of sexual partners:
    - More than one sexual partner in the last 12 months or refused to answer versus one or no sexual partners in the last 12 months (referent).
    - More than one sexual partner in the last 3 months or refused to answer versus one or no sexual partners in the last 3 months (referent).
    - At least one new sexual partners in the last 12 months or refused to answer versus no new sexual partners in the last 12 months (referent).
    - At least one new sexual partners in the last 3 months or refused to answer versus no new sexual partners in the last 3 months (referent).

### Results

#### IV. Additional demographic characteristics

**Individuals**

| Characteristic | Index mothers | | Direct offspring | | Other household members | | |
|---|---|---|---|---|---|---|---|
| | Exposed | Unexposed | Exposed | Unexposed | Exposed | Unexposed | Overall |
| n | 100 | 100 | 228 | 239 | 156 | 183 | 1,006 |
| Education, n (%) | | | | | | | |
| Below school age | 0 (0.0) | 0 (0.0) | 104 (45.6) | 112 (46.9) | 9 (5.8) | 13 (7.1) | 238 (23.7) |
| No schooling | 2 (2.0) | 0 (0.0) | 6 (2.6) | 3 (1.3) | 12 (7.7) | 4 (2.2) | 27 (2.7) |
| Any primary | 5 (5.0) | 1 (1.0) | 65 (28.5) | 72 (30.1) | 23 (14.7) | 21 (11.5) | 187 (18.6) |
| Any secondary | 73 (73.0) | 71 (71.0) | 45 (19.7) | 48 (20.1) | 60 (38.5) | 102 (55.7) | 399 (39.7) |
| Any university | 20 (20.0) | 25 (25.0) | 6 (2.6) | 0 (0.0) | 47 (30.1) | 35 (19.1) | 133 (13.2) |
| Other | 0 (0.0) | 3 (3.0) | 2 (0.9) | 4 (1.7) | 5 (3.2) | 8 (4.4) | 22 (2.2) |
| Occupation, n (%) | | | | | | | |
| No occupation | 51 (51.0) | 54 (54.0) | 31 (86.1) | 24 (96.0) | 44 (38.6) | 75 (56.4) | 279 (55.0) |
| Salaried | 14 (14.0) | 8 (8.0) | 0 (0.0) | 0 (0.0) | 35 (30.7) | 26 (19.5) | 83 (16.4) |
| Self-employed | 28 (28.0) | 26 (26.0) | 3 (8.3) | 0 (0.0) | 20 (17.5) | 21 (15.8) | 98 (19.3) |
| Works for someone else | 2 (2.0) | 0 (0.0) | 0 (0.0) | 0 (0.0) | 0 (0.0) | 0 (0.0) | 2 (0.4) |
| Student | 1 (1.0) | 2 (2.0) | 2 (5.6) | 0 (0.0) | 6 (5.3) | 4 (3.0) | 15 (3.0) |
| Other | 4 (4.0) | 10 (10.0) | 0 (0.0) | 1 (4.0) | 9 (7.9) | 7 (5.3) | 30 (5.9) |

### V. Sensitivity analyses for analyses of factors associated with HBsAg positivity.

Index mothers' antenatal recruitment HBsAg test result was used as the primary HBsAg result for index mothers and for determining household exposure status. Here we consider alternative definitions of HBsAg positivity of index mothers, to identify resolved and incident HBsAg infections, and thus alternate definitions of household exposure status:

- Enrollment
  - Index mothers: index mothers HBsAg-positive or HBsAg-negative at the single study household visit
  - Exposed direct offspring: direct offspring with an index mother who was HBsAg-positive at the single study household visit
- Either positive
  - Index mothers: index mothers HBsAg-positive at either antenatal recruitment or the single study household visit (chronic and acute infections)
  - Exposed direct offspring: direct offspring with an index mother who was HBsAg-positive at any timepoint. This subset reflects exposure to index mothers who are chronically infected (and potentially infected when the child was born) as well as exposure to recent infections of their mothers.
- Always positive
  - Index mothers: index mothers HBsAg-positive at both antenatal recruitment testing and the single study household visit (more reflective of potential chronic infection)
  - Exposed direct offspring: direct offspring with an index mother who was HBsAg-positive both timepoints, i.e. living with a mother who was likely chronically infected

**Supplementary Figure 3: Sensitivity analyses for household HBsAg prevalence**

| | PR (95% CI)* |
|---|---|
| **All household members: exposed vs unexposed** | 2.52 (0.88, 7.23) |
| **Direct offspring: exposed vs unexposed** | 3.46 (0.85, 14.01) |
| **Other: exposed vs unexposed** | 1.01 (0.24, 4.25) |
| **Exposed: direct offspring vs other** | 1.25 (0.48, 3.24) |
| **Unexposed: direct offspring vs other** | 0.91 (0.26, 3.17) |

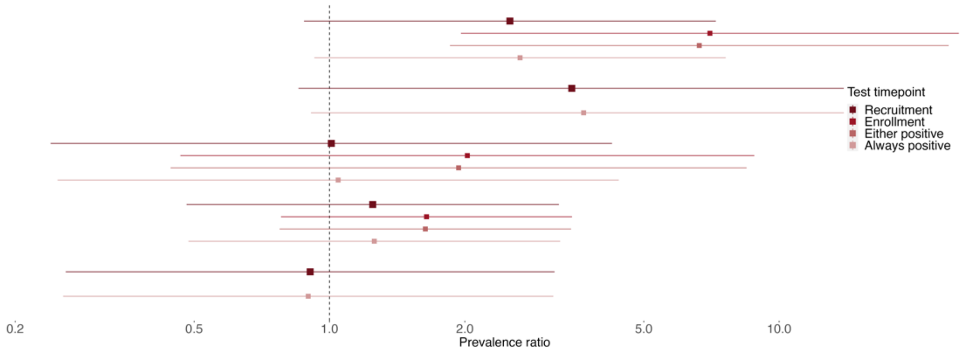


*Prevalence ratio adjusted for household clustering.

The households with male partners who were HBsAg-positive do not change with alternate definitions of exposure, and thus are not shown.

### Supplementary Figure 4: Sensitivity analyses for bivariate associations among index mothers

| | OR (95% CI)* |
|---|---|
| Age (1-year increase) | 1.06 (1.01, 1.11) |
| Marriage: Never vs married | 0.39 (0.14, 0.96) |
| Marriage: Divorced/widowed vs married | 0.78 (0.28, 2.15) |
| Upper wealth quartiles (Q4-Q2) vs lowest (Q1) | 0.73 (0.38, 1.38) |
| Shares toothbrushes in household | 1.00 (0.39, 2.55) |
| Shares razors in household | 0.67 (0.37, 1.20) |
| Shares nail clippers in household | 0.62 (0.35, 1.08) |
| Premasticates food for someone else | 0.53 (0.16, 1.60) |
| ≥1 past transfusion vs none | 1.99 (0.78, 5.50) |
| Uses street salons | 1.65 (0.89, 3.11) |
| Manicures/pedicures outside home | 1.74 (0.91, 3.41) |
| Tattoos | 0.48 (0.12, 1.58) |
| Traditional scarring | 1.00 (0.35, 2.83) |
| Has engaged in transactional sex or refused to answer vs no | 1.00 (0.53, 1.89) |
| Age of sexual debut: <18 yrs vs ≥18 yrs | 1.21 (0.66, 2.23) |
| Age of sexual debut: Refused/don't know vs ≥18 yrs | 4.53 (1.54, 16.63) |
| Sexual partners in last 3 months: ≥2 or refused vs ≤1 | 4.41 (1.35, 19.82) |
| New sexual partners in last 3 months: ≥1 or refuse vs none new | 3.63 (1.60, 9.06) |
| Sexual partners in last 12 months: ≥2 or refused vs ≤1 | 6.68 (1.76, 43.7) |
| New sexual partners in last 12 months: ≥1 or refuse vs none new | 3.32 (1.39, 8.84) |

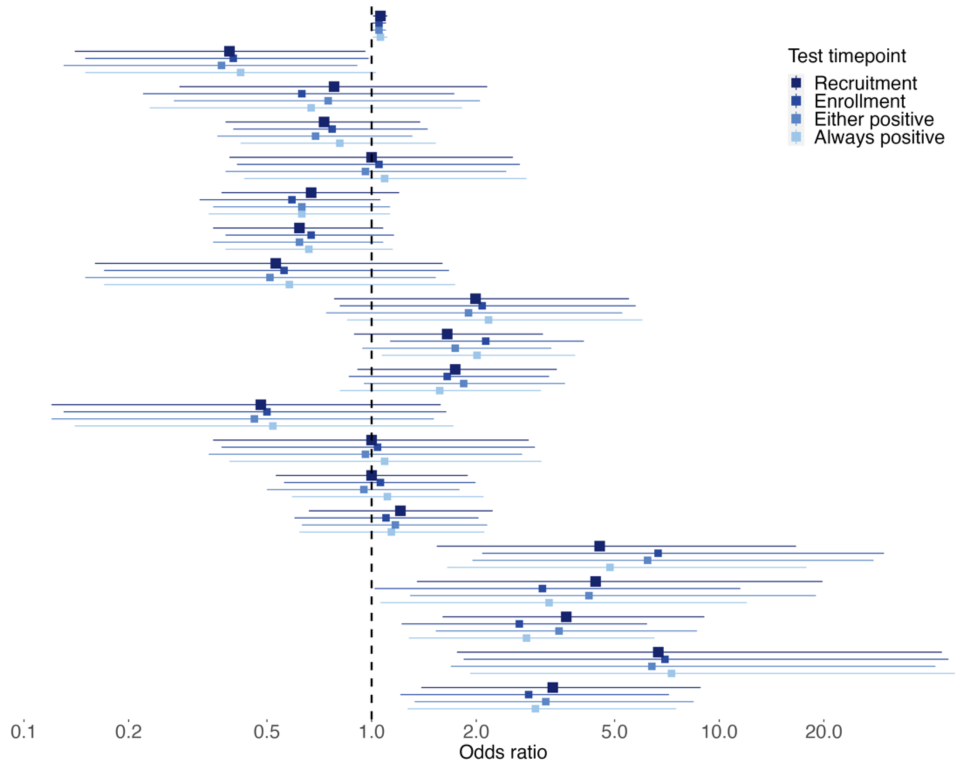


*Unadjusted odds ratio

**Supplementary Figure 5: Sensitivity analyses for bivariate associations among exposed direct offspring**

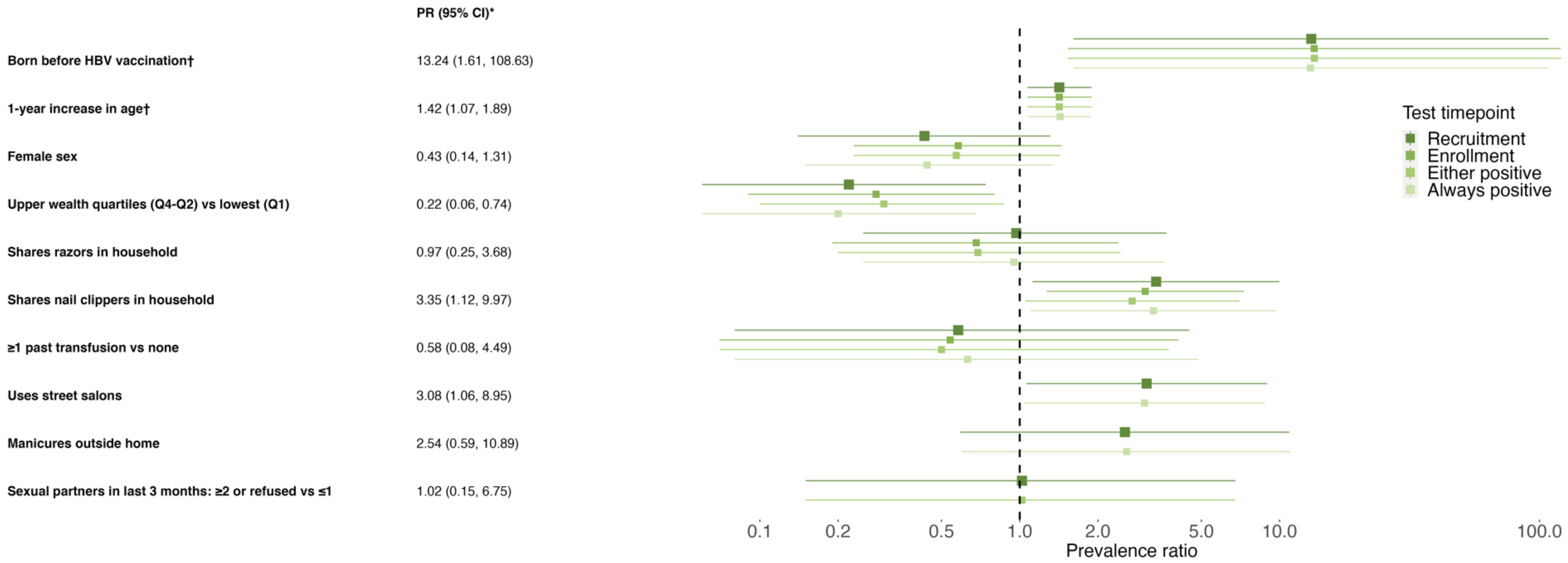


*Prevalence ratio adjusted for household clustering.
†Logistic-binomial approximation of PR due to rare outcome.
