## Supplemental Tables for "Hepatitis B virus prevalence and transmission in the households of pregnant women in Kinshasa, Democratic Republic of Congo"

**Supplemental Table 1**

| Subgroup | Scale | Subject | Variable level | Recruitment HBsAg+ (n) | Recruitment HBsAg- (n) | log(OR) | std.error | p.value | log(lower 95% CI) | log(upper 95% CI) |
|---|---|---|---|---|---|---|---|---|---|---|
| Index mothers | **Total, n** | | | **100** | **100** | | | | | |
| | Individual | Age (continuous) | 1-year increase | | | 0.054 | 0.023 | 0.021 | 0.009 | 0.101 |
| | | Marital status | Divorced/widowed | 8 | 9 | -0.243 | 0.511 | 0.635 | -1.269 | 0.766 |
| | | | Not married | 7 | 16 | -0.952 | 0.48 | 0.047 | -1.954 | -0.045 |
| | | | Married/living together | 85 | 75 | | | | | |
| | Household | Wealth | Upper 3 quartiles | 72 | 78 | -0.631 | 0.577 | 0.274 | -1.842 | 0.47 |
| | | | Lowest quartile | 28 | 22 | | | | | |
| | | Shares nail clippers in household | Shares | 41 | 53 | -0.484 | 0.285 | 0.09 | -1.048 | 0.073 |
| | | | Doesn't share | 59 | 47 | | | | | |
| | | Shares razors in household | Shares | 29 | 38 | -0.406 | 0.302 | 0.179 | -1.003 | 0.183 |
| | | | Doesn't share | 71 | 62 | | | | | |
| | | Shares toothbrushes in household | Shares | 10 | 10 | 0 | 0.471 | 1 | -0.936 | 0.936 |
| | | | Doesn't share | 90 | 90 | | | | | |
| | | Premasticated food | Yes | 5 | 9 | -0.321 | 0.328 | 0.328 | -0.972 | 0.32 |
| | | | No | 95 | 91 | | | | | |
| | Community | Past transfusions | 1+ or refused | 13 | 7 | 0.686 | 0.492 | 0.163 | -0.254 | 1.704 |
| | | | Never | 87 | 93 | | | | | |
| | | Street salon | Uses 1-4x/month or refused | 77 | 67 | 0.5 | 0.319 | 0.117 | -0.12 | 1.134 |
| | | | Doesn't use | 23 | 33 | | | | | |
| | | Manicure/pedicure | Outside home | 81 | 71 | 0.555 | 0.337 | 0.1 | -0.099 | 1.227 |
| | | | Doesn't go outside home | 19 | 29 | | | | | |
| | | Tattoo history | Has tattoos | 4 | 8 | -0.736 | 0.63 | 0.243 | -2.084 | 0.455 |
| | | | No tattoos | 96 | 92 | | | | | |
| | | Traditional scarring | Has engaged | 8 | 7 | 0.134 | 0.538 | 0.804 | -0.93 | 1.219 |
| | | | None | 92 | 92 | | | | | |
| | | Transactional sex | Has given/received money for sex | 26 | 26 | 0 | 0.322 | 1 | -0.634 | 0.634 |
| | | | Never | 74 | 74 | | | | | |
| | | Age of sexual debut | <18 years | 32 | 32 | 0.1886 | 0.3114 | 0.5448 | -0.4226 | 0.8012 |
| | | | Refused/don't know | 15 | 4 | 1.5103 | 0.5926 | 0.0108 | 0.4299 | 2.8109 |
| | | | ≥18 years | 53 | 96 | | | | | |
| | | Sexual partners in last 3 months | ≥2 or refused/don't know | 12 | 3 | 1.484 | 0.662 | 0.025 | 0.299 | 2.986 |
| | | | 1 or none | 88 | 97 | | | | | |
| | | New sexual partners in last 3 months | ≥1 new | 13 | 7 | 0.799 | 0.498 | 0.108 | -0.152 | 1.827 |
| | | | None new | 66 | 79 | | | | | |
| | | Sexual partners in last 12 months | ≥2 or refused/don't know | 12 | 2 | 1.899 | 0.777 | 0.015 | 0.565 | 3.777 |
| | | | 1 or none | 88 | 98 | | | | | |
| | | | ≥1 new or refused/don't know | 20 | 7 | 1.2 | 0.465 | 0.01 | 0.331 | 2.179 |
| | | New sexual partners in last 12 months | None new | 90 | 93 | | | | | |

**Supplemental Table 2**

| Exposure group | Subgroup | Scale | Subject | Variable level | HBsAg+, n | HBsAg-, n | Fisher's exact p-val | log(OR), adjusted for household | std.error | p.value | log(lower 95% CI) | log(upper 95% CI) |
|---|---|---|---|---|---|---|---|---|---|---|---|---|
| Exposed | Direct offspring | Total, n | | | 12 | 215 | | | | | | |
| | | Individual | Age (continuous) | 1 year increase | 12 | 215 | 0.014 (from glmer) | 0.354 | 0.145 | 0.014 | 0.07 | 0.637 |
| | | | Gender | Female | 4 | 115 | 0.24 | -0.815 | 0.874 | 0.351 | -2.528 | 0.897 |
| | | | | Male | 8 | 100 | | | | | | |
| | | | Infant vaccination (approximated) | Unvaccinated | 5 | 27 | | 2.691 | 1.184 | 0.023 | 0.371 | 5.01 |
| | | | | Vaccinated | 6 | 173 | | Ref | | | | |
| | | | Wealth | Upper 3 quartiles | 4 | 156 | 0.007 | -1.727 | 0.946 | 0.068 | -3.581 | 0.126 |
| | | Household | | Lowest quartile | 8 | 59 | | | | | | |
| | | | Shares razors, toothbrushes, or nail clippers | Yes | 9 | 113 | 0.15 | 2.391 | 1.506 | 0.112 | -0.56 | 5.342 |
| | | | | Doesn't share | 3 | 102 | | | | | | |
| | | | Premasticated food | Yes | 1 | 1 | 0.1032 | 8.898 | 5.743 | 0.121 | -2.359 | 20.155 |
| | | | | No | 11 | 214 | | | | | | |
| | | | Past transfusions | 1+ or refused | 1 | 21 | 1 | -1.48 | 1.495 | 0.322 | -4.41 | 1.451 |
| | | Community | | Never | 11 | 194 | | | | | | |
| | | | Street salon | Uses 1-4x/month or refused | 6 | 55 | 0.09 | 1.328 | 1.112 | 0.233 | -0.852 | 3.508 |
| | | | | Doesn't use | 6 | 160 | | | | | | |
| | | | Manicure/pedicure | Outside home | 2 | 23 | 0.63 | 2.218 | 1.342 | 0.098 | -0.412 | 4.848 |
| | | | | Doesn't go outside home | 10 | 192 | | | | | | |
| | | | Tattoo history | Has tattoos | 0 | 4 | 1 | -27.567 | 1.5E+07 | 1 | -30266113 | 30266058 |
| | | | | No tattoos | 12 | 211 | | | | | | |
| | | | Traditional scarring | Has engaged | 0 | 10 | 1 | -69.781 | 2.1E+07 | 1 | -41593810 | 41593671 |
| | | | | None | 12 | 205 | | | | | | |
| | | | Transactional sex | Has given/received money for sex | 1 | 0 | 0.17 | 783.991 | 6.7E+07 | 1 | -1.32E+08 | 131531740 |
| | | | | Never | 5 | 29 | | | | | | |
| | | | Age of sexual debut | <18 years | 5 | 26 | 1 | -0.093 | 5.119 | 0.986 | -10.126 | 9.94 |
| | | | | ≥18 years | 1 | 4 | | | | | | |
| | | | Sexual partners in last 3 months | ≥2 | 1 | 5 | 1 | 14.205 | 7.798 | 0.069 | -1.078 | 29.489 |
| | | | | 1 or none | 5 | 24 | | | | | | |
| | | | New sexual partners in last 3 months | ≥1 new | 1 | 1 | 1 | 34.837 | 2469.98 | 0.989 | -4806.237 | 4875.911 |
| | | | | None new | 0 | 2 | | | | | | |

| **Supplemental Table 3**<br>**Exposure** | **Subgroup** | **Scale** | **Subject** | **Variable level** | **HBsAg+, n** | **HBsAg-, n** | **Prevalence of attribute/practice** | **No Fisher's p-value reported due to low cell counts.** |
|---|---|---|---|---|---|---|---|---|
| Unexposed | Direct offspring | Total, n | | | 3 | 234 | | |
| | | Individual | Age (continuous) | 1 year increase | | | | |
| | | | Gender | Female | 2 | 139 | 1.4 | |
| | | | | Male | 1 | 95 | 1 | |
| | | | Infant vaccination (approximated) | Unvaccinated | 0 | 23 | 0 | |
| | | | | Vaccinated | 3 | 183 | 1.6 | |
| | | | Wealth | Upper 3 quartiles | 2 | 176 | 1.1 | |
| | | Household | | Lowest quartile | 1 | 58 | 1.7 | |
| | | | Shares razors, toothbrushes, or nail clip | Yes | 1 | 159 | 0.6 | |
| | | | | Doesn't share | 2 | 75 | 2.6 | |
| | | | Premasticated food | Yes | 0 | 5 | 0 | |
| | | | | No | 3 | 229 | 1.3 | |
| | | | Past transfusions | 1+ or refused | 0 | 20 | 0 | |
| | | Community | | Never | 3 | 214 | 1.4 | |
| | | | Street salon | Uses 1-4x/month or refused | 0 | 64 | 0 | |
| | | | | Doesn't use | 3 | 170 | 1.7 | |
| | | | Manicure/pedicure | Outside home | 0 | 17 | 0 | |
| | | | | Doesn't go outside home | 3 | 217 | 1.4 | |
| | | | Tattoo history | Has tattoos | 0 | 0 | 0 | |
| | | | | No tattoos | 3 | 234 | 1.3 | |
| | | | Traditional scarring | Has engaged | 0 | 8 | 0 | |
| | | | | None | 3 | 226 | 1.3 | |
| | | | Transactional sex | Has given/received money for sex | 0 | 1 | 0 | |
| | | | | Never | 0 | 22 | 0 | |
| | | | Age of sexual debut | <18 years | 0 | 23 | 0 | |
| | | | | ≥18 years | 0 | 0 | 0 | |
| | | | Sexual partners in last 3 months | ≥2 | 0 | 1 | 0 | |
| | | | | 1 or none | 0 | 22 | 0 | |
| | | | New sexual partners in last 3 months | ≥1 new | 0 | 0 | 0 | |
| | | | | None new | 0 | 1 | 0 | |

| Supplemental Table 4<br>Exposure | Subgroup | Scale | Subject | Variable level | HBsAg+, n | HBsAg-, n | Prevalence of attribute/ practice | Fisher's exact p-val |
|---|---|---|---|---|---|---|---|---|
| Exposed | Other household member | Total, n | | | 7 | 149 | | |
| | | Individual | Gender | Female | 1 | 74 | 1.3 | 0.1185 |
| | | | | Male | 6 | 75 | 7.4 | |
| | | | Marital status | Married/living | 2 | 54 | 3.6 | 0.286 |
| | | | | Not married | 5 | 40 | 11.1 | |
| | | | | Divorced/widowed | 0 | 11 | 0 | |
| | | | Infant vaccination (approximated) | Unvaccinated | 6 | 107 | 5.3 | 0.2148 |
| | | | | Vaccinated | 0 | 32 | 0 | |
| | | | Wealth | Upper 3 quartiles | 7 | 114 | 5.8 | 0.3504 |
| | | | | Lowest quartile | 0 | 35 | 0 | |
| | | Household | Shares razors, toothbrushes, or nail clippers | Yes | 5 | 90 | 5.3 | 0.7056 |
| | | | | Doesn't share | 2 | 59 | 3.3 | |
| | | | Premasticated food | Yes | 0 | 3 | 0 | 1 |
| | | | | No | 7 | 146 | 4.6 | |
| | | | Past transfusions | 1+ or refused | 1 | 8 | 11.1 | 0.3459 |
| | | | | Never | 6 | 141 | 4.1 | |
| | | Community | Street salon | Uses 1-4x/month or refused | 6 | 83 | 6.7 | 0.24 |
| | | | | Doesn't use | 1 | 66 | 1.5 | |
| | | | Manicure/pedicure | Outside home | 5 | 53 | 8.6 | 0.1025 |
| | | | | Doesn't go outside home | 2 | 96 | 2 | |
| | | | Tattoo history | Has tattoos | 0 | 8 | 0 | 1 |
| | | | | No tattoos | 7 | 141 | 4.7 | |
| | | | Traditional scarring | Has engaged | 4 | 12 | 25 | 0.0022** |
| | | | | None | 3 | 137 | 2.1 | |
| | | | Transactional sex | Has given/received money for sex | 1 | 23 | 4.2 | 1 |
| | | | | Never | 6 | 86 | 6.5 | |
| | | | Age of sexual debut | <18 years | 6 | 75 | 7.4 | 0.6731 |
| | | | | ≥18 years | 1 | 34 | 2.9 | |
| | | | Sexual partners in last 3 months | ≥2 | 0 | 33 | 0 | 0.1891 |
| | | | | 1 or none | 7 | 76 | 8.4 | |
| | | | New sexual partners in last 3 months | ≥1 new | 1 | 18 | 5.3 | 1 |
| | | | | None new | 3 | 35 | 7.9 | |

**Supplemental Table 5**

| Exposure | Subgroup | Scale | Subject | Variable level | HBsAg+, n | HBsAg-, n | Prevalence of attribute/ practice | Fisher's exact p-val |
|---|---|---|---|---|---|---|---|---|
| Unexposed | Other household member | Total, n | | | 5 | 178 | | |
| | | Individual | Gender | Female | 3 | 98 | 3 | 1 |
| | | | | Male | 2 | 80 | 2.4 | |
| | | | Marital status | Married/living | 1 | 48 | 2 | 0.501 |
| | | | | Not married | 1 | 65 | 1.5 | |
| | | | | Divorced/widowed | 1 | 15 | 6.3 | |
| | | | Infant vaccination (approximated) | Unvaccinated | 1 | 16 | 5.9 | 0.5551 |
| | | | | Vaccinated | 3 | 29 | 9.4 | |
| | | | Wealth | Upper 3 quartiles | 5 | 149 | 3.2 | 1 |
| | | | | Lowest quartile | 0 | 29 | 0 | |
| | | Household | Shares razors, toothbrushes, or nail clippers | Yes | 4 | 122 | 3.2 | 1 |
| | | | | Doesn't share | 1 | 56 | 1.8 | |
| | | | Premasticated food | Yes | 0 | 3 | 0 | 1 |
| | | | | No | 5 | 175 | 2.8 | |
| | | | Past transfusions | 1+ or refused | 0 | 21 | 0 | 1 |
| | | | | Never | 5 | 157 | 3.1 | |
| | | Community | Street salon | Uses 1-4x/month or refused | 3 | 95 | 3.1 | 1 |
| | | | | Doesn't use | 2 | 83 | 2.4 | |
| | | | Manicure/pedicure | Outside home | 1 | 69 | 1.4 | 0.6599 |
| | | | | Doesn't go outside home | 4 | 108 | 3.6 | |
| | | | Tattoo history | Has tattoos | 0 | 7 | 0 | 1 |
| | | | | No tattoos | 5 | 171 | 2.8 | |
| | | | Traditional scarring | Has engaged | 0 | 26 | 0 | 1 |
| | | | | None | 5 | 152 | 3.2 | |
| | | | Transactional sex | Has given/received money for sex | 1 | 25 | 3.8 | 0.488 |
| | | | | Never | 2 | 103 | 1.9 | |
| | | | Age of sexual debut | <18 years | 2 | 94 | 2.1 | 1 |
| | | | | ≥18 years | 1 | 34 | 2.9 | |
| | | | Sexual partners in last 3 months | ≥2 | 0 | 21 | 0 | 1 |
| | | | | 1 or none | 3 | 106 | 2.8 | |
| | | | New sexual partners in last 3 months | ≥1 new | 0 | 12 | 0 | 1 |
| | | | | None new | 1 | 46 | 2.1 | |
